# Women with poor brain health at time of ischemic stroke endure worse outcomes compared to men

**DOI:** 10.64898/2026.02.21.26346614

**Authors:** Kenda Alhadid, Erik Lindgren, Robert W. Regenhardt, Arne G Lindgren, Christina Jern, Jane MacGuire, Natalia S. Rost, Markus D. Schirmer, the MRI-GENIE and GISCOME Investigators

**Affiliations:** Department of Neurology, Mass General Brigham, Harvard Medical School, Boston, MA, USA; J. Philip Kistler Stroke Research Center, Massachusetts General Hospital, Harvard Medical School, Boston, MA, USA; Department of Clinical Neuroscience, Institute of Neuroscience and Physiology, Sahlgrenska Academy, University of Gothenburg; Department of Neurology, Sahlgrenska University Hospital, Region Västra Götaland, Gothenburg, Sweden; Department of Neurology, UTHealth Houston Neurosciences, Houston, TX, USA; Department of Neurology, Skåne University Hospital; Department of Clinical Sciences Lund, Neurology, Lund University, Lund, Sweden; Department of Laboratory Medicine, Institute of Biomedicine, Sahlgrenska Academy, University of Gothenburg; Department of Clinical Genetics and Genomics, Sahlgrenska University Hospital, Region Västra Götaland, Gothenburg, Sweden; University of Technology Sydney, Sydney, Australia

**Author notes:** Correspondence: Kenda Alhadid, 175 Cambridge Street, Boston, MA, USA 02114.

## Abstract

**Importance:** Prognosticating functional independence after an acute stroke is critical for anticipatory guidance and rehabilitation planning. Here we demonstrate that poor brain health at the time of incident stroke is linked to worse functional outcomes for women compared to men.

**Objective:** To determine if brain health at time of stroke presentation has a differential effect on functional outcomes between men and women.

**Design:** Retrospective cross-sectional study.

**Setting:** Analysis conducted in 2025 with multi-center patient data that included participants from two large acute ischemic stroke cohorts; local (GASROS) and multinational (MRI-GENIE) between the years 2003 and 2011.

**Participants:** Clinical data collected for enrolled study participants included demographic data, medical history of hypertension, diabetes mellitus, hyperlipidemia, smoking status, acute stroke severity as measured by National Institutes of Health Stroke Scale (NIHSS), stroke etiology, and modified Rankin Scale (mRS) score at 90 days post-stroke. Brain health was quantified as effective reserve derived from acute neuroimaging data.

**Exposure(s):** designated sex, retrieved from registration records.

**Main Outcome:** Functional outcome was measured by mRS scores at 90 days post-stroke, in men and women with poor, moderate, or good brain health at time of stroke injury.

**Results:** A total of 1039 patients were included in the analysis, 37.8 % women, median age 67 [interquartile range 56-77]. Women with poor brain health (i.e. lowest quartile of effective reserve) had worse functional outcomes at 90 days (55.6% with mRS>2) compared to men with poor brain health (31.2% with mRS>2: p < 0.001). This difference between men and women was not observed in categories of moderate or good brain health. There was no observed significant difference in stroke severity, volume of acute lesion, burden of white matter hyperintensities, or stroke etiology between men and women with poor brain health.

**Conclusions and Relevance:** Brain health at the time of incident stroke has a differential effect on functional outcomes at 90 days between men and women. Women with poor brain health endure disproportionately worse outcomes compared to men. This highlights an important step in understanding sex-specific vulnerability in early recovery post-stroke, and can inform disposition, rehabilitation services, and resource allocation planning.

## INTRODUCTION

Stroke is the leading cause of serious long term disability in the United States and significantly impacts mobility in more than half of survivors over the age of 65 ^1^. Globally, neurological disorders are the leading cause of disability adjusted life years (DALYs) and stroke has the highest age standardized DALYs of all disorders ^2^. The vast majority of strokes are due to ischemic injury (∼87%) ^3^. Recent data shows that stroke incidence is increasing in young and midlife adults over the past few decades ^4^, with women being disproportionately affected ^5^.

Many studies have been dedicated to understanding predictors of outcome after incident stroke ^6^. Not only is this information valuable for acute management (such as disposition from hospital admission), but also for facilitating rehabilitation efforts and counseling survivors. It has been established that clinical measures of overall health such as existing comorbidities and functional status at the time of stroke, in addition to a multitude of socioeconomic factors, all majorly impact stroke recovery. Their interactions with general health, and specifically brain health, are profound and complex ^7,8^. Brain health is defined as attaining and maintaining optimal neurological functioning in the face of aging and confers resilience to neurological injury ^9^. Special attention has been given to the impact of vascular risk factors ^10^ and cardiac disease ^11^ on brain health.

Sex-specific differences in stroke patients have been described in all aspects of stroke including but not limited to: clinical presentation, accuracy and timeliness of diagnosis, and management ^12,13^. Differences between men and women have also been observed in the diagnosis of stroke, incidence rates across age groups, impact of vascular risk factors on stroke incidence, etiology of stroke, and various aspects of outcomes post-stroke ^14–16^. With regard to functional outcomes, data have shown that women present with more severe stroke syndromes and have worse outcomes compared to men even after correcting for older age at stroke diagnosis, stroke lesion volume, and baseline functional status ^17–19^. Recent data shows that when examining specific age groups, post-stroke mortality was higher in women aged 50-70 compared to men ^20^. However, despite the observed sex differences, we continue to have a relatively limited understanding of the complex processes that drive these observed effects of biological sex on stroke and its neurological sequelae ^21^.

Recent works have leveraged large datasets with clinical and neuroimaging data, to study observed sex-specific differences in outcomes post stroke. Techniques such as lesion-symptom analysis, stroke topography analysis, structural disconnection measures, and quantification of white matter hyperintensity (WMH) volumes at time of stroke, have all shown sex-specific implications for recovery after ischemic stroke ^22–26^. We recently developed and expanded the concept of effective reserve (eR), a latent variable neuroimaging biomarker of brain health that improves stroke outcome modeling by incorporating three key variables: age, brain volume, and burden of white matter hyperintensities on standard brain MRI scans ^27–29^. We have also found that eR outperformed other neuroimaging biomarkers and clinical markers alone in modeling stroke outcome ^30^. However, the effects of sex-specific differences in brain health for functional outcome remain unknown.

In this study, we evaluated whether brain health at the time of stroke had a differential effect on functional outcomes at 90 days post-stroke between men and women. Clinical and neuroimaging data were accessed for two large ischemic stroke patient cohorts with standard-of-care neuroimaging sequences. Utilizing the acute imaging data for estimating eR, we evaluated sex differences in 90-day post-stroke outcome, measured by modified Rankin Scale (mRS), stratified by low, medium, or high levels of brain health at the time of incident stroke.

## METHODS

### Study Cohorts and Patient Characteristics

Two acute ischemic stroke (AIS) patient cohorts were studied: Patients admitted to Massachusetts General Hospital (MGH) and enrolled in the Genes Associated with Stroke Risk and Outcomes Study (GASROS: model development cohort), and the MRI-GENetics Interface Exploration (MRI-GENIE: model validation cohort), an international, multi-site and hospital-based cohort of AIS patients ^31^. Inclusion criteria for patients in both cohorts were: I) age > 18 years and II) confirmed neuroimaging diagnosis of AIS within 48 hours of admission. Clinical data regarding stroke severity, vascular risk factors, and functional outcomes (as measured by the mRS at 90 days post-stroke extracted from patient or caregiver interview or based on review of clinical assessment) were retrieved for all study participants. Institutional review board (IRB) approval at MGH was obtained for all subjects enrolled locally at MGH (2001P001186), as was approval from local ethics committee/IRB for each site that enrolled patients in the MRI-GENIE study.

### Study design and neuroimaging data

The study design was retrospective and cross-sectional. Two MRI sequences were studied for each subject: diffusion-weighted imaging (DWI) and T2 fluid-attenuated inversion recovery (T2-FLAIR) sequences acquired as standard-of-care imaging at each center within 48 hours from clinical presentation. Parameters of MRI data acquisition in the GASROS cohort were as follows: T2 FLAIR imaging (TR 5000ms, TE 62-116ms, TI 2200 ms, FOV 220-240mm), and DWI (single-shot echo-planar imaging; 1-5 B0 volumes, 6-30 diffusion directions with b=1000 s/mm^2^, 1-3 averaged volumes). Imaging data parameters from MRI-GENIE are described in detail elsewhere ^31^.

Stroke lesion volume was manually calculated from acute diffusion sequences for GASROS and automatically quantified for MRI-GENIE participants ^32^. Volume of white matter hyperintensities and brain volume were automatically computed from T2-FLAIR sequences using analysis pipelines developed for acute stroke imaging ^28,33^, and used in the estimation of eR scores that were categorized into three groups of brain health (Fig. 1).

### Data analysis

Quality of brain, stroke lesion, and WMH segmentations were manually reviewed in a quality control step by visual inspection. Volumes were calculated by multiplying the number of voxels within each mask by voxel size. Lesion load and WMH load were defined as the ratio of lesion and WMH volume to brain volume, respectively, and logit transformed for normality. Age was reported in decades, and brain volume in dm^3^. eR was formulated as a latent variable for every study subject, given by eR ∼ Age + WMH load + Brain volume, with coefficients used as reported in our prior study ^27^.

Stroke pathogenetic mechanisms as reported by TOAST criteria ^34^ and cardiovascular risk factors were compared between men and women from both cohorts. eR scores were categorized into three groups based on quartiles (poor brain health: first/lowest quartile of eR scores; moderate brain health: second and third quartiles; and good brain health: fourth quartile) derived from the GASROS cohort and evaluated on the MRI-GENIE cohort. Functional outcome distributions within quartiles, in addition to stroke lesion load and WMH lesion load, were compared between women and men across the three categories using the Kolmogorov-Smirnov test. All analyses were performed in R ^35^ with significance set at p<0.05.

## RESULTS

A total of 1039 patients were included in the analysis. In comparison between men and women included in the analysis, there was no significant difference in stroke severity or in stroke etiology based on the TOAST classification. Men had significantly higher rates of smoking and hypertension compared to women (Table 1). No significant differences were observed between cohorts in patient age or percentage of women and men included. Patient characteristics, stratified by cohort, are presented in Supplementary Table 1.

**Table 1:**
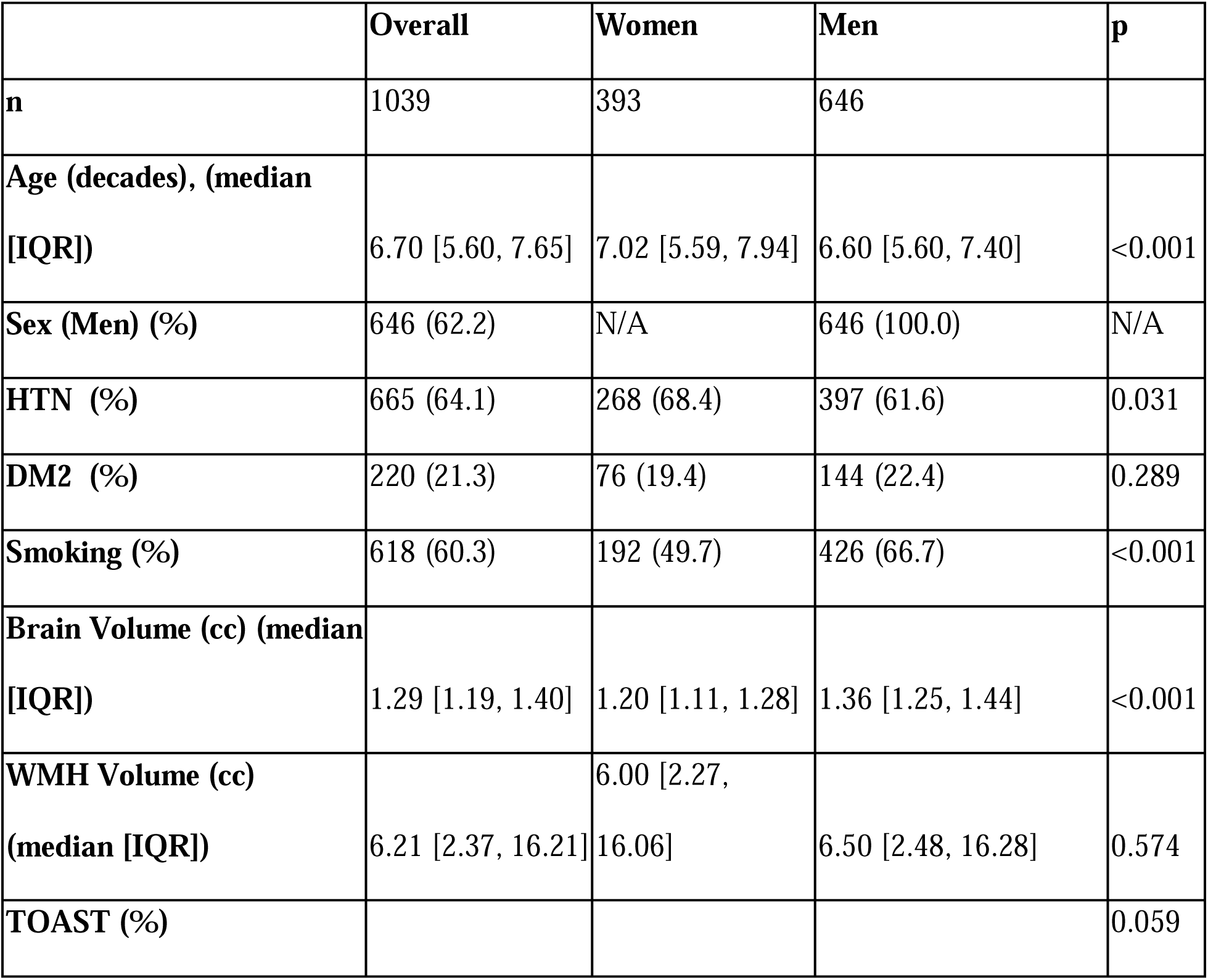
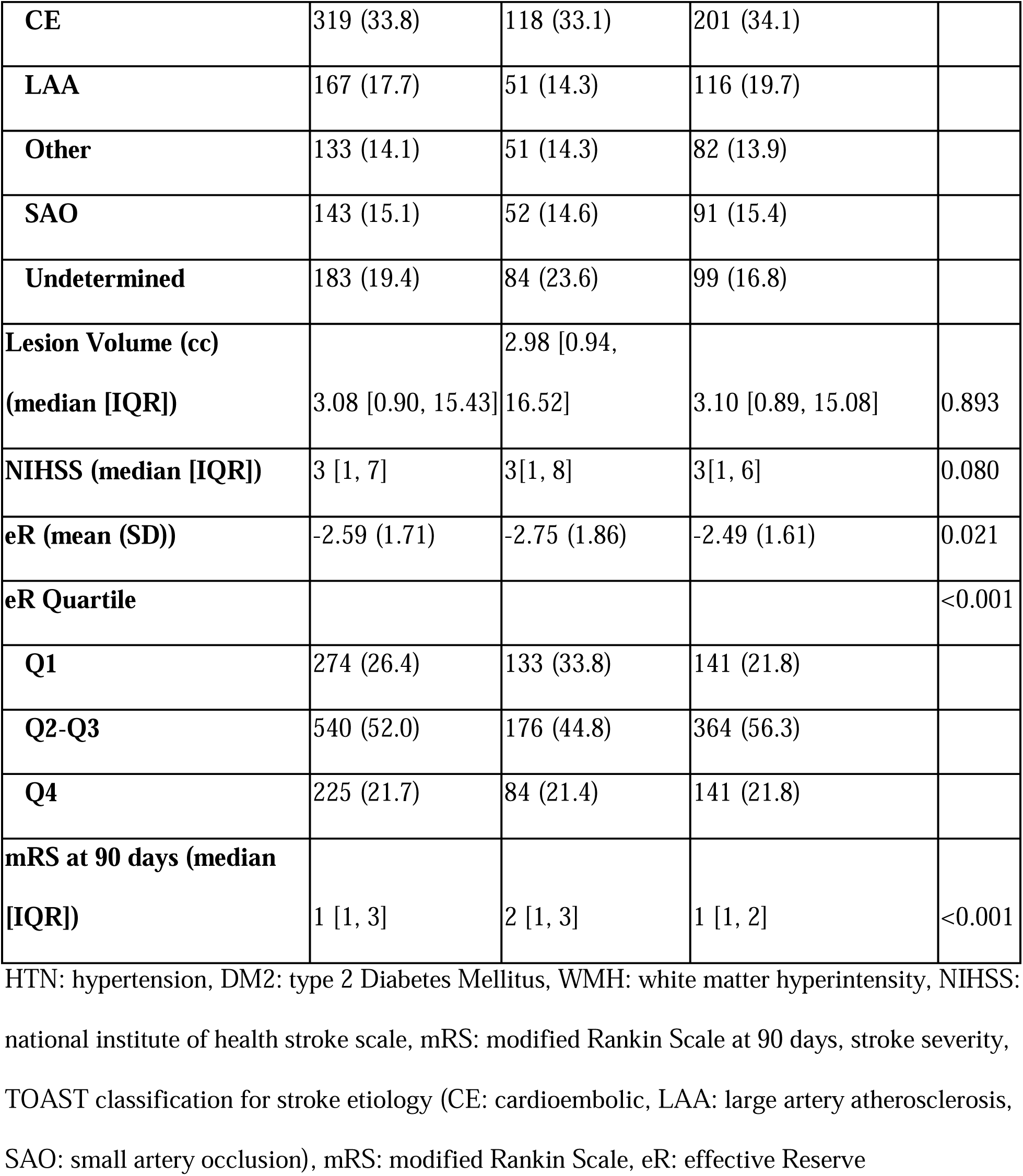
Cohort characteristics of the combined MRI-GENIE and GASROS studies and comparison between men and women included in this study.

Women with evidence of poor brain health at the time of incident stroke (eR values in the lowest quartile) had significantly worse functional outcomes at 90 days post-stroke compared to men counterparts ( p < 0.001; Fig. 2), with a significant proportion with an mRS score of 3 or higher [55.6% vs 31.2% for women vs. men (Fisher’s exact test p < 0.001)].

There were no significant differences in functional outcomes between men and women in the moderate (eR values in second and third quartile) or good brain health (eR values in fourth quartile) categories, nor difference in age between men and women for each mRS outcome. Across all categories of brain health, men and women had comparable WMH and stroke lesion burden, while women had significantly smaller brain volumes (Supplementary Table 2).

## DISCUSSION

In this study we evaluated whether brain health at the time of stroke impacts functional outcomes in a differential manner between the sexes. We report that poor brain health at the time of stroke assessed on routine clinical imaging, has significant implications regarding outcomes for women compared to men at 90 days post-stroke. A significantly larger proportion of women with poor brain health (55.6%) are found to be functionally dependent at 90 days (mRS of 3 or higher) compared to only (31.2%) of men in the same category of brain health.

Across all categories of brain health, women had significantly smaller total brain volumes, but no significant differences between men and women in stroke lesion load or white matter hyperintensity load were found. This observed difference in brain volume is likely due to known anthropomorphic differences. Brain volume is an important determinant of functional outcomes after ischemic stroke, with a larger brain implying a better functional outcome ^33,36^. However, this alone is unlikely to explain the differences in outcome between men and women within the category of low brain health as women’s brains were significantly smaller in volume in all categories of brain health compared to men, with no observed effect on outcome.

Outcome prognostication in the acute phase of stroke injury is typically informed by clinical variables and neuroimaging findings such as acute lesion volume. Neuroimaging biomarkers can, and have been, leveraged to inform outcome predictions ^37^. Prior studies have utilized these to explain why women endure worse outcomes compared to men after ischemic stroke, and the majority focused on features relevant to the stroke injury itself such as lesion size, lesion location, connected structures, and stroke etiology ^22,38^. Specific imaging findings that have been found to be related to poor functional outcome, include severity of pre-existing white matter disease ^39^, with more recent data showing that white matter disease pattern, corticospinal tract involvement, and left parieto-occipital lobe infarcts, all have sex-specific implications for recovery ^23,40,41^. While these studies mostly examined imaging features specific to the stroke injury itself such as lesion volume and location, we evaluated the impact of brain health at the time of injury on recovery. Previous work demonstrated that including brain health in prediction models of stroke outcome improves model performance ^27^, and here we demonstrate that for women specifically, poor brain health purveys a worse functional outcome. We argue that eR, as an imaging biomarker of brain health, encompasses sex-specific differences that should be accounted for in outcome modeling.

Possible explanations for this finding include differences related to traditional cardiovascular risk profiles, individual differences in exposures, and genetic susceptibilities that are not typically captured in most studies. Women experience higher rates of depression and more disabling higher-order cognitive impairments post-stroke ^42,43^ ^40,41^ that could impact recovery in the subacute phase, and could also explain the observation that they are more likely to be offered palliative care with comfort measures and less likely to be offered rehabilitation services compared to men ^44^. Cohort data for most acute stroke studies do not typically include reproductive history, or adverse pregnancy outcomes (APOs), nor history of exposure to hormonal therapy. These sex-specific clinical data include factors that impact cerebrovascular health, change the structure of the white matter and gray matter, and alter brain volume ^45–47^. Furthermore, traditional cardiovascular risk factors such as diabetes mellitus, hypertension, and obesity have been shown to have sex-specific differences ^48^ throughout the lifetime not just in the biology of the disease but in how it is managed and how it interacts with cardiovascular and cerebrovascular health over time ^49^. Obesity, which is more prevalent in women, has a stronger association with stroke risk than that observed in men ^12^ and imaging studies have shown that obesity produces changes in structural neuroimaging biomarkers of brain health with a likely sex-specific interaction ^50,51^. Notably, even a history of obesity in early life confers a higher risk of ischemic stroke in women independently of their later BMI ^52^.

There are some limitations in this study. There is no treatment or rehabilitation data available for the study subjects included in the analysis. However, both cohorts included patients with relatively mild to moderate stroke severity as measured by NIHSS at the time of presentation, and enrollment of participants was prior to mechanical thrombectomy becoming a standard of care therapeutic intervention. Thrombolytic therapy was likely offered universally to those who met clinical criteria for treatment, however, future studies with treatment information available are warranted. While various standardized clinical scores aim to ascertain functional independence and ambulatory capacity post-stroke, no single measure can fully characterize outcome. Nonetheless, mRS is widely used in clinical trials to ascertain outcomes, and revealed sex-specific differences in post-stroke recovery in earlier studies that showed women endured worse post-stroke outcomes ^44,52^. Lastly, there are numerous clinical and socioeconomic factors that impact functional outcomes in stroke survivors that are not uniformly collected in acute stroke cohorts such as post-stroke depression, access to care and resources, amongst other factors that can impact recovery. Future studies with detailed prospective collection of these variables are needed to characterize their contributions to brain health and functional outcomes post-stroke.

Strengths of our study include the relative homogeneity of our study subjects and their diagnosis; enrolled prospectively in acute stroke cohorts with clinical and acute neuroimaging obtained uniformly per standard-of-care clinical practice, minimizing selection bias. We employed automated segmentation pipelines developed specifically for acute stroke imaging datasets, manually inspected output for quality control, and then disease burden size relative to brain volume was computed, allowing for the best possible estimation of effect of stroke lesion size and WMH burden. Comprehensive clinical data was retrieved with data regarding pre-existing cardiovascular risk factors extracted, to ascertain whether past medical history influenced observed differences in outcomes between men and women with poor brain health. In summary, we show that brain health at the time of incident stroke has implications for functional outcomes with a differential effect between men and women, with poor brain health imparting worse functional outcomes for women. Neuroimaging markers of brain health should be included in stroke recovery models.

## Data Availability

Data will be made available without investigator support for purposes of replicating the results, after approval of a submitted data request form and obtaining the relevant local IRBs approval, and with a signed data-use agreement.

---

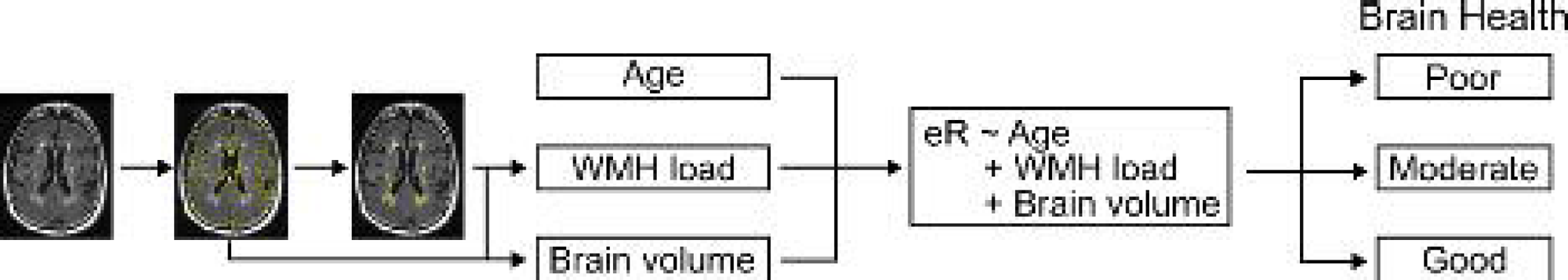

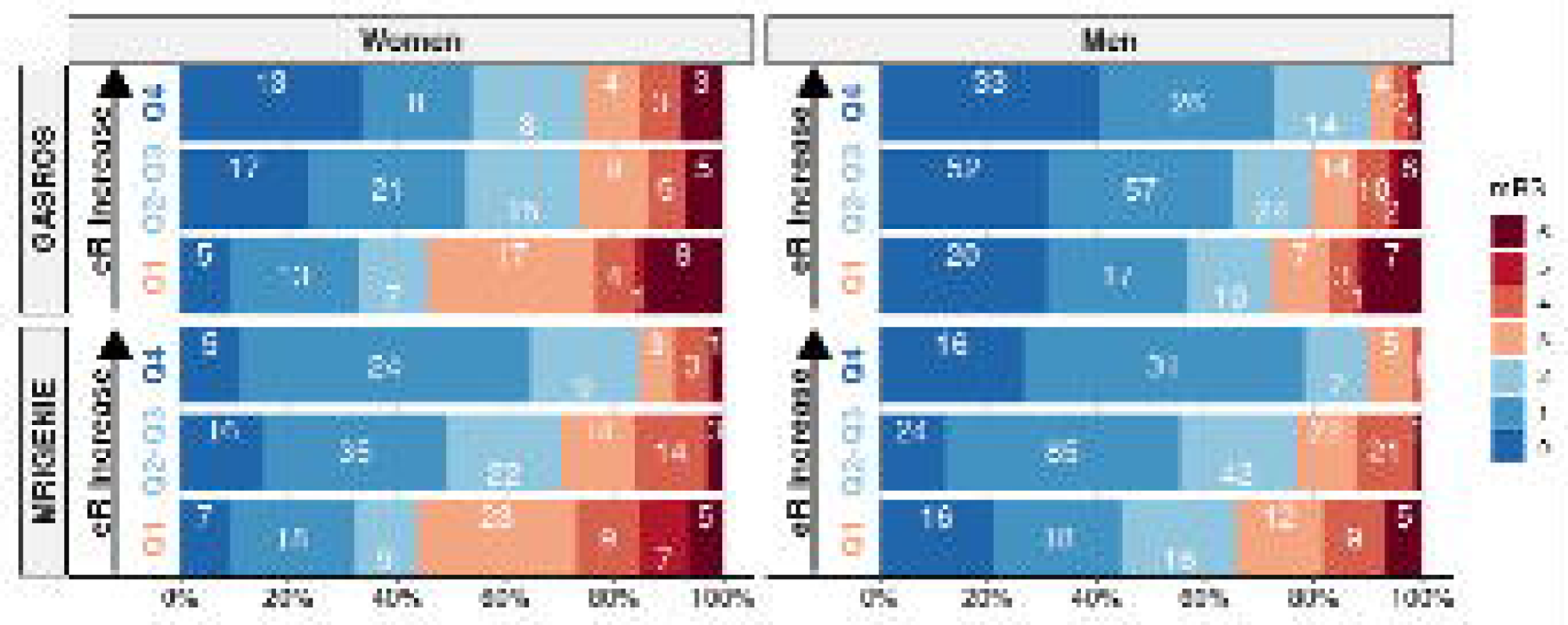

## Notes

### Competing Interest Statement

The authors have declared no competing interest.

### Funding Statement

This study did not receive any funding.

### Author Declarations

Institutional review board (IRB) approval at Massachusetts General Hospital was obtained for all subjects enrolled locally at MGH (2001P001186), as was approval from local ethics committee/IRB for each site that enrolled patients in the MRI-GENIE [MRI-GENetics Interface Exploration] study. Datasets were de-identified prior to use in this study.

## REFERENCES

1. Tsao CW, Aday AW, Almarzooq ZI, et al. Heart Disease and Stroke Statistics—2023 Update: A Report From the American Heart Association. Circulation. 2023;147(8):e93–e621. doi:10.1161/CIR.0000000000001123

2. GBD 2021 Nervous System Disorders Collaborators. Global, regional, and national burden of disorders affecting the nervous system, 1990-2021: a systematic analysis for the Global Burden of Disease Study 2021. Lancet Neurol. 2024;23(4):344–381. doi:10.1016/S1474-4422(24)00038-3

3. Andersen KK, Olsen TS, Dehlendorff C, Kammersgaard LP. Hemorrhagic and ischemic strokes compared: stroke severity, mortality, and risk factors. Stroke. 2009;40(6):2068–2072. doi:10.1161/STROKEAHA.108.540112

4. Ekker MS, Verhoeven JI, Vaartjes I, van Nieuwenhuizen KM, Klijn CJM, de Leeuw FE. Stroke incidence in young adults according to age, subtype, sex, and time trends. Neurology. 2019;92(21):e2444–e2454. doi:10.1212/WNL.0000000000007533

5. Leppert MH, Burke JF, Lisabeth LD, et al. Systematic Review of Sex Differences in Ischemic Strokes Among Young Adults: Are Young Women Disproportionately at Risk? Stroke. 2022;53(2):319–327. doi:10.1161/STROKEAHA.121.037117

6. Matsumoto K, Nohara Y, Soejima H, Yonehara T, Nakashima N, Kamouchi M. Stroke Prognostic Scores and Data-Driven Prediction of Clinical Outcomes After Acute Ischemic Stroke. Stroke. 2020;51(5):1477–1483. doi:10.1161/STROKEAHA.119.027300

7. Hackman DA, Farah MJ, Meaney MJ. Socioeconomic status and the brain: mechanistic insights from human and animal research. Nat Rev Neurosci. 2010;11(9):651–659. doi:10.1038/nrn2897

8. Nguyen MTH, Sakamoto Y, Maeda T, et al. Influence of Socioeconomic Status on Functional Outcomes After Stroke: A Systematic Review and Meta Analysis. J Am Heart Assoc. 2024;13(9):e033078. doi:10.1161/JAHA.123.033078

9. Rost NS, Salinas J, Jordan JT, et al. The Brain Health Imperative in the 21st Century-A Call to Action: The AAN Brain Health Platform and Position Statement. Neurology. 2023;101(13):570–579. doi:10.1212/WNL.0000000000207739

10. Gorelick PB, Furie KL, Iadecola C, et al. Defining Optimal Brain Health in Adults: A Presidential Advisory From the American Heart Association/American Stroke Association. Stroke. 2017;48(10):e284–e303. doi:10.1161/STR.0000000000000148

11. Testai FD, Gorelick PB, Chuang PY, et al. Cardiac Contributions to Brain Health: A Scientific Statement From the American Heart Association. Stroke. 2024;55(12):e425–e438. doi:10.1161/STR.0000000000000476

12. Rexrode KM, Madsen TE, Yu AYX, Carcel C, Lichtman JH, Miller EC. The Impact of Sex and Gender on Stroke. Circ Res. 2022;130(4):512–528. doi:10.1161/CIRCRESAHA.121.319915

13. Yu AYX, Penn AM, Lesperance ML, et al. Sex Differences in Presentation and Outcome After an Acute Transient or Minor Neurologic Event. JAMA Neurol. 2019;76(8):962–968. doi:10.1001/jamaneurol.2019.1305

14. Di Carlo A, Lamassa M, Baldereschi M, et al. Sex differences in the clinical presentation, resource use, and 3-month outcome of acute stroke in Europe: data from a multicenter multinational hospital-based registry. Stroke. 2003;34(5):1114–1119. doi:10.1161/01.STR.0000068410.07397.D7

15. Reeves MJ, Bushnell CD, Howard G, et al. Sex differences in stroke: epidemiology, clinical presentation, medical care, and outcomes. Lancet Neurol. 2008;7(10):915–926. doi:10.1016/S1474-4422(08)70193-5

16. Phan HT, Blizzard CL, Reeves MJ, et al. Factors contributing to sex differences in functional outcomes and participation after stroke. Neurology. 2018;90(22):e1945–e1953. doi:10.1212/WNL.0000000000005602

17. Dehlendorff C, Andersen KK, Olsen TS. Sex Disparities in Stroke: Women Have More Severe Strokes but Better Survival Than Men. J Am Heart Assoc. 2015;4(7):e001967. doi:10.1161/JAHA.115.001967

18. Manwani B, McCullough LD. On the Basis of Sex: Outcomes After Ischemic Stroke. Stroke. 2019;50(9):2285–2287. doi:10.1161/STROKEAHA.119.025955

19. Silva GS, Lima FO, Camargo ECS, et al. Gender differences in outcomes after ischemic stroke: role of ischemic lesion volume and intracranial large-artery occlusion. Cerebrovasc Dis Basel Switz. 2010;30(5):470–475. doi:10.1159/000317088

20. Yu AYX, Austin PC, Rashid M, et al. Sex Differences in Intensity of Care and Outcomes After Acute Ischemic Stroke Across the Age Continuum. Neurology. 2023;100(2):e163–e171. doi:10.1212/WNL.0000000000201372

21. Bonkhoff AK, Coughlan G, Perosa V, et al. Sex differences in age-associated neurological diseases—A roadmap for reliable and high-yield research. Sci Adv. 2025;11(10):eadt9243. doi:10.1126/sciadv.adt9243

22. Bonkhoff AK, Bretzner M, Hong S, et al. Sex-specific lesion pattern of functional outcomes after stroke. Brain Commun. 2022;4(2):fcac020. doi:10.1093/braincomms/fcac020

23. Bonkhoff AK, Schirmer MD, Bretzner M, et al. Outcome after acute ischemic stroke is linked to sex-specific lesion patterns. Nat Commun. 2021;12(1):3289. doi:10.1038/s41467-021-23492-3

24. Bonkhoff AK, Schirmer MD, Bretzner M, et al. The relevance of rich club regions for functional outcome post-stroke is enhanced in women. Hum Brain Mapp. 2023;44(4):1579–1592. doi:10.1002/hbm.26159

25. Etherton MR, Wu O, Cougo P, et al. Structural Integrity of Normal Appearing White Matter and Sex-Specific Outcomes After Acute Ischemic Stroke. Stroke. 2017;48(12):3387–3389. doi:10.1161/STROKEAHA.117.019258

26. Lohner V, Pehlivan G, Sanroma G, et al. Relation Between Sex, Menopause, and White Matter Hyperintensities. Neurology. 2022;99(9):e935–e943. doi:10.1212/WNL.0000000000200782

27. Schirmer MD, Etherton MR, Dalca AV, et al. Effective reserve: a latent variable to improve outcome prediction in stroke. J Stroke Cerebrovasc Dis Off J Natl Stroke Assoc. 2019;28(1):63–69. doi:10.1016/j.jstrokecerebrovasdis.2018.09.003

28. Schirmer MD, Giese AK, Fotiadis P, et al. Spatial Signature of White Matter Hyperintensities in Stroke Patients. Front Neurol. 2019;10. doi:10.3389/fneur.2019.00208

29. Schirmer, Alhadid, Kenda, Regenhardt, Robert W., Rost, Natalia S. Quantifying brain health in acute ischemic stroke through effective reserve. medRxiv. Published online March 2024.

30. Lindgren E, Angeleri L, Bretzner M, et al. Magnetic Resonance Imaging Markers of Brain Health Improve Assessment of Functional Outcome After Acute Ischemic Stroke: A Quantitative Comparison Study. medRxiv. Preprint posted online May 3, 2025:2025.05.02.25326881. doi:10.1101/2025.05.02.25326881

31. Giese AK, Schirmer MD, Donahue KL, et al. Design and rationale for examining neuroimaging genetics in ischemic stroke: The MRI-GENIE study. Neurol Genet. 2017;3(5):e180. doi:10.1212/NXG.0000000000000180

32. Wu O, Winzeck S, Giese AK, et al. Big Data Approaches to Phenotyping Acute Ischemic Stroke Using Automated Lesion Segmentation of Multi-Center Magnetic Resonance Imaging Data. Stroke. 2019;50(7):1734–1741. doi:10.1161/STROKEAHA.119.025373

33. Alhadid K, Regenhardt RW, Rost NS, Schirmer MD. Brain volume is a better biomarker of outcomes in ischemic stroke compared to brain atrophy. Front Stroke. 2024;3. doi:10.3389/fstro.2024.1468772

34. Adams HP, Bendixen BH, Kappelle LJ, et al. Classification of subtype of acute ischemic stroke. Definitions for use in a multicenter clinical trial. TOAST. Trial of Org 10172 in Acute Stroke Treatment. Stroke. 1993;24(1):35–41. doi:10.1161/01.str.24.1.35

35. R Core Team. R: A language and environment for statistical computing. Published online 2024.

36. Schirmer MD, Donahue KL, Nardin MJ, et al. Brain Volume: An Important Determinant of Functional Outcome After Acute Ischemic Stroke. Mayo Clin Proc. 2020;95(5):955–965. doi:10.1016/j.mayocp.2020.01.027

37. Heiss WD, Kidwell CS. Imaging for Prediction of Functional Outcome and Assessment of Recovery in Ischemic Stroke. Stroke J Cereb Circ. 2014;45(4):1195–1201. doi:10.1161/STROKEAHA.113.003611

38. Ryu WS, Chung J, Schellingerhout D, et al. Biological Mechanism of Sex Difference in Stroke Manifestation and Outcomes. Neurology. 2023;100(24):e2490–e2503. doi:10.1212/WNL.0000000000207346

39. Kissela B, Lindsell CJ, Kleindorfer D, et al. Clinical prediction of functional outcome after ischemic stroke: the surprising importance of periventricular white matter disease and race. Stroke. 2009;40(2):530–536. doi:10.1161/STROKEAHA.108.521906

40. Ryu WS, Chung J, Schellingerhout D, et al. Biological Mechanism of Sex Difference in Stroke Manifestation and Outcomes. Neurology. 2023;100(24):e2490–e2503. doi:10.1212/WNL.0000000000207346

41. Lindenberg R, Renga V, Zhu LL, Betzler F, Alsop D, Schlaug G. Structural integrity of corticospinal motor fibers predicts motor impairment in chronic stroke. Neurology. 2010;74(4):280–287. doi:10.1212/WNL.0b013e3181ccc6d9

42. Guo X, Phan C, Batarseh S, Wei M, Dye J. Risk factors and predictive markers of post-stroke cognitive decline–A mini review. Front Aging Neurosci. 2024;16. doi:10.3389/fnagi.2024.1359792

43. Kim JS, Lee KB, Roh H, Ahn MY, Hwang HW. Gender Differences in the Functional Recovery after Acute Stroke. J Clin Neurol Seoul Korea. 2010;6(4):183–188. doi:10.3988/jcn.2010.6.4.183

44. Miller EC, Conley P, Alirezaei M, et al. Associations between adverse pregnancy outcomes and cognitive impairment and dementia: a systematic review and meta-analysis. Lancet Healthy Longev. 2024;5(12):100660. doi:10.1016/j.lanhl.2024.100660

45. Kantarci K, Tosakulwong N, Lesnick TG, et al. Effects of hormone therapy on brain structure. Neurology. 2016;87(9):887–896. doi:10.1212/WNL.0000000000002970

46. Miller EC, Kauko A, Tom SE, et al. Risk of Midlife Stroke After Adverse Pregnancy Outcomes: The FinnGen Study. Stroke. 2023;54(7):1798–1805. doi:10.1161/STROKEAHA.123.043052

47. Rajendran A, Minhas AS, Kazzi B, et al. Sex-specific differences in cardiovascular risk factors and implications for cardiovascular disease prevention in women. Atherosclerosis. 2023;384:117269. doi:10.1016/j.atherosclerosis.2023.117269

48. Regensteiner JG, Reusch JEB. Sex Differences in Cardiovascular Consequences of Hypertension, Obesity, and Diabetes. J Am Coll Cardiol. 2022;79(15):1492–1505. doi:10.1016/j.jacc.2022.02.010

49. Gazdzinski S, Kornak J, Weiner MW, Meyerhoff DJ. Body Mass Index and Magnetic Resonance Markers of Brain Integrity in Adults. Ann Neurol. 2008;63(5):652–657. doi:10.1002/ana.21377

50. Kroll DS, Feldman DE, Biesecker CL, et al. Neuroimaging of Sex/Gender Differences in Obesity: A Review of Structure, Function, and Neurotransmission. Nutrients. 2020;12(7):1942. doi:10.3390/nu12071942

51. Mikkola U, Rissanen I, Kivelä M, et al. Overweight in Adolescence and Young Adulthood in Association With Adult Cerebrovascular Disease: The NFBC1966 Study. Stroke. 2024;55(7):1857–1865. doi:10.1161/STROKEAHA.123.045444

52. Gargano JW, Reeves MJ, Paul Coverdell National Acute Stroke Registry Michigan Prototype Investigators. Sex differences in stroke recovery and stroke-specific quality of life: results from a statewide stroke registry. Stroke. 2007;38(9):2541–2548. doi:10.1161/STROKEAHA.107.485482

