## Supplemental Tables 1 and 2 for "Women with poor brain health at time of ischemic stroke endure worse outcomes compared to men"

**SUPPLEMENTAL DATA:**

Supplemental Table 1. Patient characteristics of study participants. HTN: hypertension, DM2: Type

2 Diabetes Mellitus, NIHSS: National Institute of Health Stroke Scale, TOAST: Trial of OG 10172

in Acute Stroke Treatment, CE: cardioembolic stroke, LAA: Large artery atherosclerosis, SAO:

small artery occlusion, Other: other determined cause, Undetermined cause), WMH: white matter

hyperintensities)

|  | **Overall** | **GASROS** | **MRI-GENIE** | **p** |
| --- | --- | --- | --- | --- |
| **n** | 1039 | 479 | 560 |  |
| **Age (decades) (median [IQR])** | 6.70 [5.60, 7.65] | 6.58 [5.50, 7.63] | 6.80 [5.70, 7.70] | 0.108 |
| **Sex (Men; %)** | 646 (62.2) | 313 (65.3) | 333 (59.5) | 0.06 |
| **HTN (%)** | 665 (64.1) | 295 (61.6) | 370 (66.3) | 0.13 |
| **DM2 (%)** | 220 (21.3) | 96 (20.0) | 124 (22.4) | 0.401 |
| **Smoking (%)** | 618 (60.3) | 317 (66.2) | 301 (55.1) | <0.001 |
| **NIHSS** | 3 [1, 7] | 3 [1, 6] | 4 [2, 7] | <0.001 |
| **TOAST (%)** |  |  |  | <0.001 |
| **CE** | 319 (33.8) | 177 (38.6) | 142 (29.2) |  |
| **LAA** | 167 (17.7) | 78 (17.0) | 89 (18.3) |  |
| **Other** | 133 (14.1) | 111 (24.2) | 22 (4.5) |  |
| **SAO** | 143 (15.1) | 57 (12.4) | 86 (17.7) |  |
| **Undetermined** | 183 (19.4) | 36 (7.8) | 147 (30.2) |  |
| **mRS (median [IQR])** | 1.00 [1.00, 3.00] | 1.00 [0.00, 2.00] | 1.00 [1.00, 3.00] | 0.001 |
| **Brain Volume (median [IQR])** | 1.29 [1.19, 1.40] | 1.31 [1.19, 1.41] | 1.28 [1.18, 1.39] | 0.022 |
| **WMH Load (median [IQR])** | -5.30 [-6.34, -4.32] | -5.33 [-6.44, -4.29] | -5.26 [-6.21, -4.35] | 0.47 |
| **Stroke Lesion Load (median [IQR])** | -6.00 [-7.27, -4.40] | -6.37 [-7.75, -4.60] | -5.70 [-6.85, -4.28] | <0.001 |

Supplemental table 2. Brain Volume (1000 cc) , Stroke lesion load, and White Matter hyperintensity load comparisons between men and women in quartiles of eR. Q1 = poor brain health, Q2-3 = moderate brain health, Q4 good brain health

|  | **Quartiles of eR (or brain health)** | **N**  **Women** | **N**  **Men** | **Mean (+/- SD)**  **Women** | **Mean (+/- SD)**  **Men** | **p** |
| --- | --- | --- | --- | --- | --- | --- |
| **Brain Volume (dm^3^)** | Q1(poor) | 134 | 140 | 1.15 ± 0.13 | 1.26 ± 0.12 | <0.001 |
|  | Q2-Q3(moderate) | 176 | 364 | 1.20 ± 0.12 | 1.35 ± 0.13 | <0.001 |
|  | Q4 (good) | 83 | 141 | 1.27 ± 0.11 | 1.42 ± 0.11 | <0.001 |
| **Lesion Load** | Q1 | 56 | 64 | -6.34 ± 2.03 | -6.65 ± 1.91 | 0.402 |
|  | Q2-Q3 | 71 | 167 | -6.69 ± 1.81 | -6.27 ± 2.02 | 0.112 |
|  | Q4 | 39 | 81 | -5.55 ± 2.21 | -5.75 ± 1.92 | 0.628 |
| **WMH Load** | Q1 | 134 | 140 | -4.18 ± 0.68 | -4.08 ± 0.67 | 0.222 |
|  | Q2-Q3 | 176 | 364 | -5.45 ± 0.95 | -5.31 ± 0.97 | 0.112 |
|  | Q4 | 83 | 141 | -7.11 ± 0.96 | -7.03 ± 0.98 | 0.561 |
